# Transmission Dynamics and Control Methodology of COVID-19: a Modeling Study

**DOI:** 10.1101/2020.03.29.20047118

**Authors:** Hongjun Zhu, Yan Li, Xuelian Jin, Jiangping Huang, Xin Liu, Ying Qian, Jindong Tan

## Abstract

The coronavirus disease 2019 (COVID-19) has grown up to be a pandemic within a short span of time. To investigate transmission dynamics and then determine control methodology, we took epidemic in Wuhan as a study case. Unfortunately, to our best knowledge, the existing models are based on the common assumption that the total population follows a homogeneous spatial distribution, which is not the case for the prevalence occurred both in the community and in hospital due to the difference in the contact rate. To solve this problem, we propose a novel epidemic model called SEIR-HC, which is a novel epidemic model with two different social circles. Using the model alongside the exclusive optimization algorithm, the spread process of COVID-19 epidemic in Wuhan city is reproduced and then the propagation characteristics and unknown data are estimated. Furthermore, the control measures implemented in Wuhan are assessed and the control methodology of COVID-19 is discussed to provide guidance for limiting the epidemic spread.

## 1. Introduction

A total of 113,702 confirmed cases have been reported worldwide as of 10:00 CET on Mar. 10. The statistical data shows that the COVID-19 outbreak constitutes an enormous threat to human in the world. Many factors, including human connectivity, urbanization, as well as international travel, present global challenges for disease prevention and its spread control [4]. Fortunately, mathematical models offer valuable tools for understanding epidemiological patterns and then for decision-making in global health.

However, modeling the transmission dynamics of epidemic is still a challenging task. A prime cause is the difficulty in obtaining the reliable data for the required data is patchy, delayed, and even wrong [1, 2, 5, 6]. The second cause is that the epidemic transmission scarcely occurs in the absence of interventions. Life activities and travel, which are tightly linked to the spread of infection, make the case more complex. The last but not least reason is that there are too many incidences, which cannot be ignored anyhow, happened in hospital where the contact rate is entirely different from that in the community. All of those pose difficulties for the general epidemic models to work. Additional problem encountered here is related to optimal estimation of the parameters of the epidemic model [8].

In order to probe the propagation characteristics and transmission mechanism, several attempts have been done on modeling the transmission dynamics of the COVID-19, using observations of the index case in Wuhan[2, 5-7, 9]. Unfortunately, the results are different and even contradictory. Moreover, there is no comparison of model output with real-world observations. Such a comparison is necessary to establish the performance of the models. In addition, the models fail to consider the difference of contact rate between in communities and in hospitals. This may limit the accuracy of the proposed models and hence the reliability of results.

To resolve this problem, this paper presents a novel epidemic model, which based on the standard assumption: the population is divided into susceptible (S), exposed (E), infectious (I), and recovered (R) groups[11]. The model proposed here, referred to as SEIR-HC for simplicity, extends the susceptible–exposed-infectious–removed (SEIR) model to handle epidemic transmission in two different but coupled social circles. Moreover, a two-step optimization is built for the parameter estimation.

Furthermore, with the limited data, transmission process of COVID-19 in Wuhan city reproduced by the proposed model. Then, the propagation characteristics and unreported data are estimated, and the end time of COVID-19 is also predicted. In the end, the control measures implemented in Wuhan are assessed and the control methodology of COVID-19 is discussed to provide guidance to cease the COVID-19 outbreak.

The remainder of the paper is structured as follows: Section 2 introduces the previous work; Section 3 defines the related terminology; Section 4 explains the SEIR-HC model in detail; Section 5 describes the two-step optimization for parameter estimation; Section 6 shows the analysis results, and finally, Section 7 states the conclusions.

## 2. Previous work

Transmission dynamics of an epidemic disease have long been a topic of active research. In 1866, Farr[12] mathematically described the cattle-plague epidemic by means of curve fitting method. In 1889, P. D. En’ko presented a discrete-time model in which the population consists of infectious individuals and susceptible individuals[13]. In 1906, Hamer[14] proposed that the incidence depends on the product of the densities of the susceptibles and infectives. The susceptible–infectious–removed (SIR) model was formulated by Martini in 1921[15, 16]. In 1927, Kermack and McKendrick [17] investigated the SIR model in a homogeneous population by using differential equations and discovered that the epidemic course would inevitably be terminated once the density of population becomes smaller than a critical threshold. Stone et al.[18] analyzed the rationale of the pulse vaccination strategy with the SIR model.

The SIR model is ideally suited to describe the process of virus spread. However, it is not always congruent with epidemic course. Some infections do not confer any long lasting immunity. In this case, one should resort to the SIS model, in which the infectious becomes susceptible again, rather than recovered. The SIS model has many potential applications such as modeling the spread behavior of computer viruses[19]. On the other hand, if the epidemic lasts for a very long time, births and deaths heavily affect the population size. So, Hethcote [20] considered births and deaths in the deterministic models of SIS and SIR.

On account of the fact that the SIS and SIR model only hold for the case without a latent period, which is not the case for many kinds of infectious diseases. To alleviate this problem, Cooke[21] proposed an epidemic model for the case that after a fixed time the infected susceptibles can get infectious. In this model, the population is divided into four classes of individuals: susceptibles, exposed individuals, infectives and recovered individuals. Such an epidemic model is known as the SEIR model. Later, Longini[22] presented a general formulation of the discrete-time epidemic model with permanent immunity. Lipsitch et al.[23] estimated the infectiousness of SARS and likelihood of an outbreak with modified SEIR model. Krylova and Earn[16] found that the dynamical structures of SEIR models have a less effect on the stage duration distribution, relative to that of the SIR models.

During the disease spread, a considerable degree of chance enters into the conditions under which fresh infections take place. Therefore, statistical fluctuations should be taken into account for a more precise analysis[24]. For this reason, Bailey[25] introduced probability distribution into the SIS epidemic model. In the standard models, the incubation and infectious periods are typically assumed to be exponential distribution, which makes the model sensitive to stochastic fluctuations[11, 26]. Because of more robustness, Weibull distribution is used to investigate severe acute respiratory syndrome by Marc Lipsitch et al [23].

For the stochastic case, even the simplest representations present difficulties in obtaining algebraic solutions[8]. To address this issue, Saunders[27] constructed an approximate maximum likelihood estimator for the chain SEIR model by using the Poisson approximation to the Binomial distribution. See [13] and[28] for an extensive review of classic epidemic models.

Indeed, a family of SEIR models is being used to support epidemic control, elimination, and eradication efforts. Recently, Wu et al. [7] provided an estimate of the size of the COVID-19 epidemic based on the SEIR model. Using an assumption of Poisson-distributed daily time increments, Read et al.[9] fitted a deterministic SEIR model to the daily number of confirmed cases in Chinese cities and cases reported in other countries/regions. Zhao et al.[6] estimated the number of unreported cases in mainland China based on the assumption that the initial growth phase followed an exponential growth pattern.

To sum up, a number of models have been proposed to formulate the transmission process of epidemic disease, which lay a good foundation for our work. Unfortunately, none of these studies have been done on the epidemic transmission under two different but coupled conditions. A mathematical model, relating to the processes, is desired to explore, anticipate, understand, and predict the effects of feedbacks within such complex systems, including changes caused by intervention.

## 3. Terminology

In many cases, canonical representation will be the starting point for obtaining a clear or concise expression. For this reason, let us introduce the notation used throughout before describing our work. Fig. 1 presents a pictorial display of disease natural history into order to make these terms easy to be understood.

**Fig. 1.**
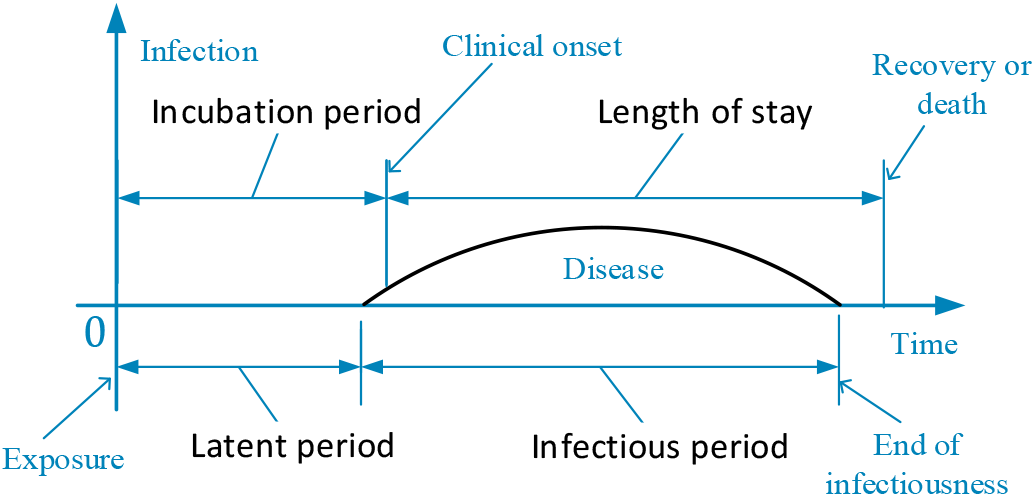
Disease natural history.

1. latent period: time from exposure to onset of infectiousness, during which the infectious agent develops in the vector, and at the end of which the vector can infect a susceptible individuals[21];
2. Incubation period: time from exposure to first appearance of clinical symptoms of infection[28];
3. Infectious period: period during which an infected person can transmit a pathogen to a susceptible[7];
4. Length of stay: time from the day of admission in the hospital to the day of discharge, i.e., the number of days a patient stayed in a hospital for treatment[29];
5. Serial interval: time from the onset of symptoms in an index case to the onset of symptoms in a subsequent case infected by the index patient [23];
6. Susceptibles: individuals who have possibility of contacting with infectious individuals but still stay uninfected[17];
7. Exposed individuals: individuals in the latent period, who are infected but not yet infectious[28];
8. Infectives: individuals who are infectious in the sense that they are capable of transmitting the infection[28];
9. Removed individuals: individuals who are removed by recovery and death, among them recovers obtain permanent infection-acquired immunity[17, 28];
10. Quarantined individuals: suspected or exposed individuals who are separated and controlled to see if they become sick[23];
11. Isolated individuals: infectives who are separated and controlled to avoid disease transmission[23];
12. Basic reproduction number: expected number of secondary infectious cases generated by an average infectious case in an entirely susceptible population[23];
13. Contact rate: average number of individuals with which one infective have an adequate contact in unit time, where an adequate contact is an interaction which results in infection [28, 30];
14. Incidence rate: rate of new infections[31], or, more precisely, total number of exposed individuals who move into infective class in unit time;
15. Death rate: the death probability of a person per day on average [17].

## 4. SEIR-HC epidemic model

The basic principle is that if the model is tightly close to the real world, then the optimization algorithm will converge to the most reasonable solution. Based on this idea, we build a new SEIR model with intervention mechanism which takes two different social circles into consideration.

### 4.1 Assumptions

The usefulness of a mathematical model depends on the existence of its solution[8]. For real world applications, a mathematical model of a complex physical situation tends to involve a certain amount of simplification. However, a suitable simplification is not straightforward to obtain a balance between simplicity and practicality. To resolve this dilemma, generality of epidemic and particularity of COVID-19 are considered simultaneously and then a set of assumptions are determined carefully as follows.

1. The population is homogeneous and uniformly mixed [21, 30, 32].
2. Recovered individuals are permanently immune and newborn infants have temporary passive immunity to the infection [28].
3. Infectiousness remains constant during an infectious period[28].
4. The natural disease-independent death rate is constant throughout the population[26].
5. The disease-caused death rate is a time-independent constant.
6. Latent period and infectious period follow Weibull distribution[23], which is a versatile distribution that has the ability to take on the characteristics of other types of distribution.
7. Contact rate is constant over the entire infectious period [22].
8. The first index case is infected on Dec 1, 2019 [7].
9. Travel behavior was not affected by disease before lockdown on Jan 23,2020[7].

### 4.2 Symbols

Consider a time interval (*t, t* + *h*], where *h* represents the length between the time points at which measurements are taken, here h = 1 day. For convenience, a variable *X* at a time interval (*t, t* + 1] is represented as *X*_(*t*)_. Then, the variables and parameters are denoted as follows:

*S*_(*t*)_ : number of susceptibles at time *t*;

*E*_(*τ,t*)_: number of exposed individuals at time *t* who were first infected at time *t* − *τ*;

*E*_(*t*)_: total number of exposed individuals at time *t*;

*I*_(*τ,t*)_: number of infectives at time t who were first at time *t* − *τ*;

*I*_(*t*)_: total number of infectives at time *t*;

*R*_(*t*)_: number of removed individuals at time *t*;

*Z*_(*t*)_ : number of infectives which is equivalent to force of infection of wildlife in the Huanan

Seafood Wholesale Market;

*N*_(*t*)_: size of population at time *t*, that is, total number of susceptibles, exposed individuals, infectives and removed individuals at time *t*;

*C*_*in*(*t*)_: number of inbound travellers every day in Wuhan at time *t*;

*C*_*t*(*t*)_: number of outbound travellers every day in Wuhan at time *t*;

*α*_*c*_: contact rate in the community;

*α*_*h*_: contact rate in hospitals;

*β*_(*τ*)_: incidence rate of exposed individuals who infected *τ* days ago, which follow the Weibull distribution;

*k*_*β*_: shape parameter of the Weibull probability density function (PDF) of incidence rate *β*;

*λ*_*β*_: scale parameter of the Weibull PDF of the incidence rate *β*;

*γ*_(*τ*)_: removal rate of infectives by disease-caused death or recovery who have been infectious for *τ* days, which follow the Weibull distribution;

*k*_*γ*_: shape parameter of the Weibull PDF of the removal rate *γ*;

*λ*_*γ*_: scale parameter of the Weibull PDF of the removal rate *γ*;

*ξ*: proportion of hospitalized infectives to total number of infectives;

*ζ*: proportion of quarantined susceptibles to total number of susceptibles;

*P*_*E*_: maximum of latent period;

*P*_*I*_: maximum of infectious period;

*v*: disease-independent death rate.

For notational convenience, index c is used to denote community and h hospital in the following expressions. The variables without the indices c and h mean they are applicable in both cases.

### 4.3 Formulating SEIR-HC epidemic model

From classical SEIR model, the population is roughly classified as four classes: susceptible, exposed, infectious, and recovered individuals. Among them, exposed and infectious individuals fall into a series of groups according to disease progression in our work so that the Weibull distribution, which armed with high generalization capability[33], can match accurately number of individuals and duration time.

In order to accommodate the quarantine and isolation measures, the standard SEIR structure was modified, as shown in Fig. 2. Note that the size of population is varied with control measures and may be affected by the inbound and outbound travellers.

**Fig. 2.**
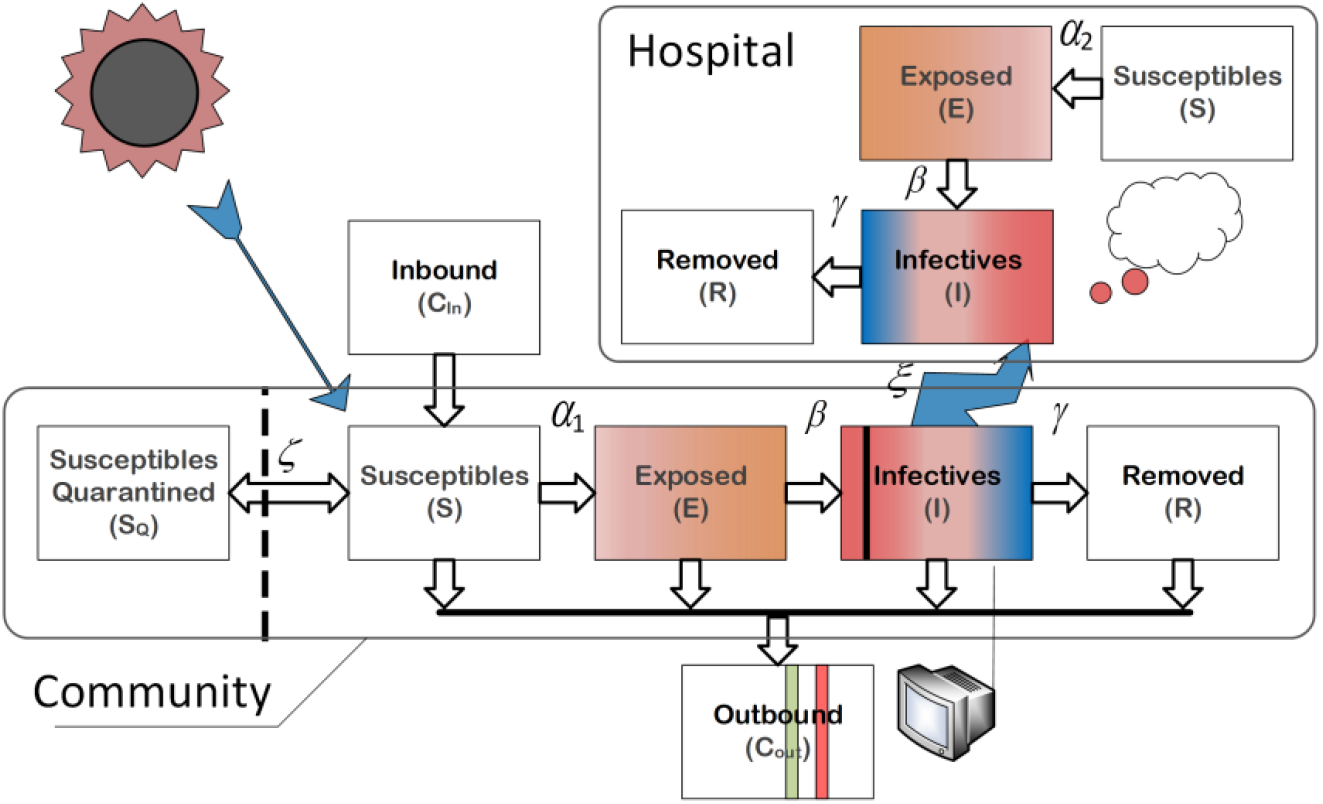
SEIR-HC model for COVID-19 transmission in Wuhan city.

For the individuals in hospital and community, disease transmissions are different in infection pattern but share the common features of the virus. For this reason, two populations are analyzed separately for the sake of accuracy. With the reservations mentioned in Section 4.1, we used the SEIR-HC model to simulate the epidemic process in Wuhan. From the rules of node dissemination, the dynamic transfer equations of the SEIR-HC model are stated as follows.

For the individuals in the community,

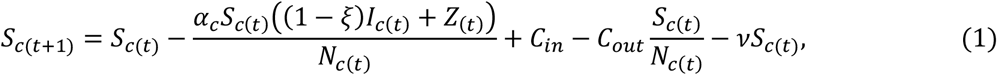

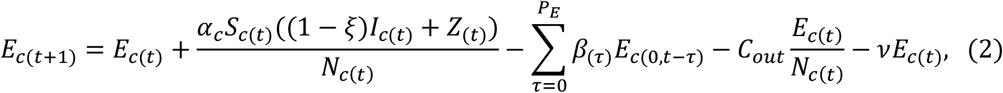

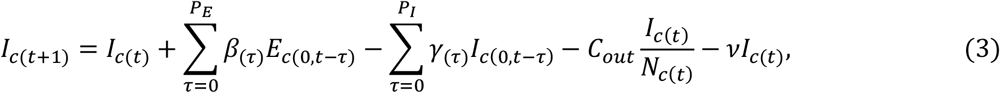

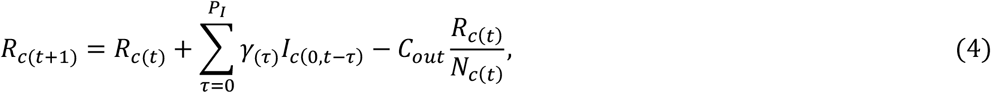

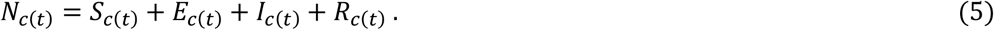

Given contact rate *α, αS*_(*t*)_/*N*_(*t*)_ is the average number of susceptibles with which per infective contacts in unit time, and thus *αS*_(*t*)_*I*_(*t*)_/*N*_(*t*)_ is the total number of susceptibles with which *I*_(*t*)_ infectives contacts in unit time (i.e., the total number of new infections in unit time). Notice that the population of Wuhan city was dramatically reduced from 14 million to 9 million before lockdown on the morning of Jan 23, 2020. On the day, the preventive and control measures of category A infectious diseases were implemented to fight against COVID-19. Therefore, we assume that *N*_*c*(*t*)_, *S*_*c*(*t*)_, *E*_*c*(*t*)_, *I*_*c*(*t*)_, *R*_*c*(*t*)_ decrease proportionately by 5/14 and then the number of susceptibles became 5*ζN*_*c*(*t*)_/14 for restrictions on outdoor activities on the same day. We further assume that the disease-independent death rate *v* is 1.50959 × 10^−5^, which is the same as that reported by the Wuhan government in December 2019[34]. According to the data presented by Wu et al[7], *C*_*in*_, *C*_*out*_ are set to 490,856, 505,646 before Jan 10, 814,046, 720,859 from Jan 11 to Jan 22, and 0, 0 after Jan 22.

For the individuals in hospital,

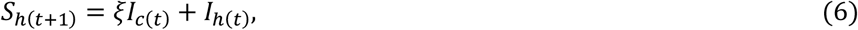

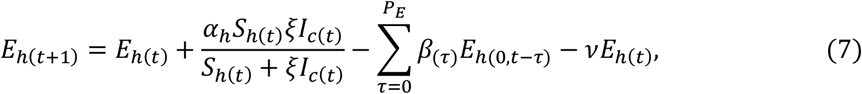

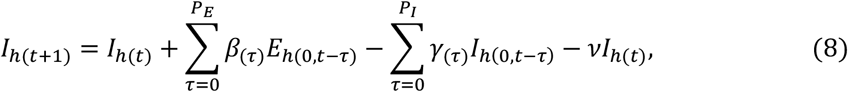

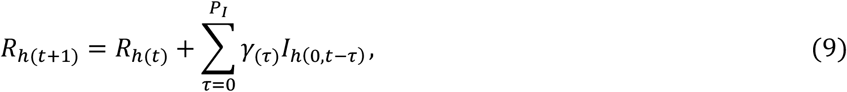

where

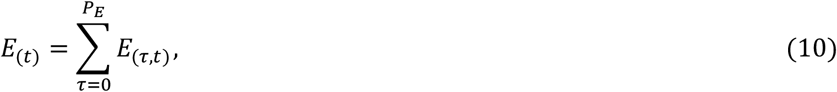

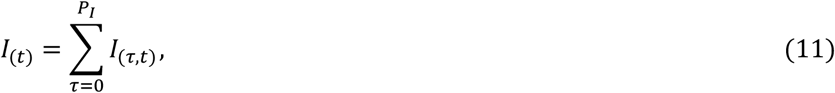

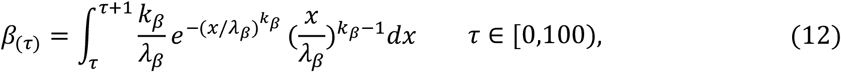

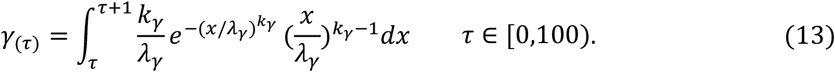

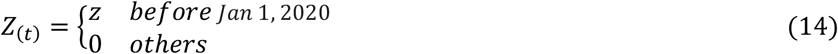

Depending on the data provided by Li et al (in Fig. 3), there are no more than 5 new cases every day from Dec 8 to Dec 28. At the same time, the basic reproduction number of the COVID-19 in Wuhan must be more than 1 or else the outbreak is impossible[23, 35]. In addition, a susceptible can be infected within 15 seconds of standing next to an infective. For these reasons, it is highly probable that the parameter z, which reflects the force of infection of the Huanan Seafood Market, is small and hence let z=1. Obviously, if let *ξ* = 0, then SEIR-HC is easy to degrade into the SEIR model.

**Fig. 3.**
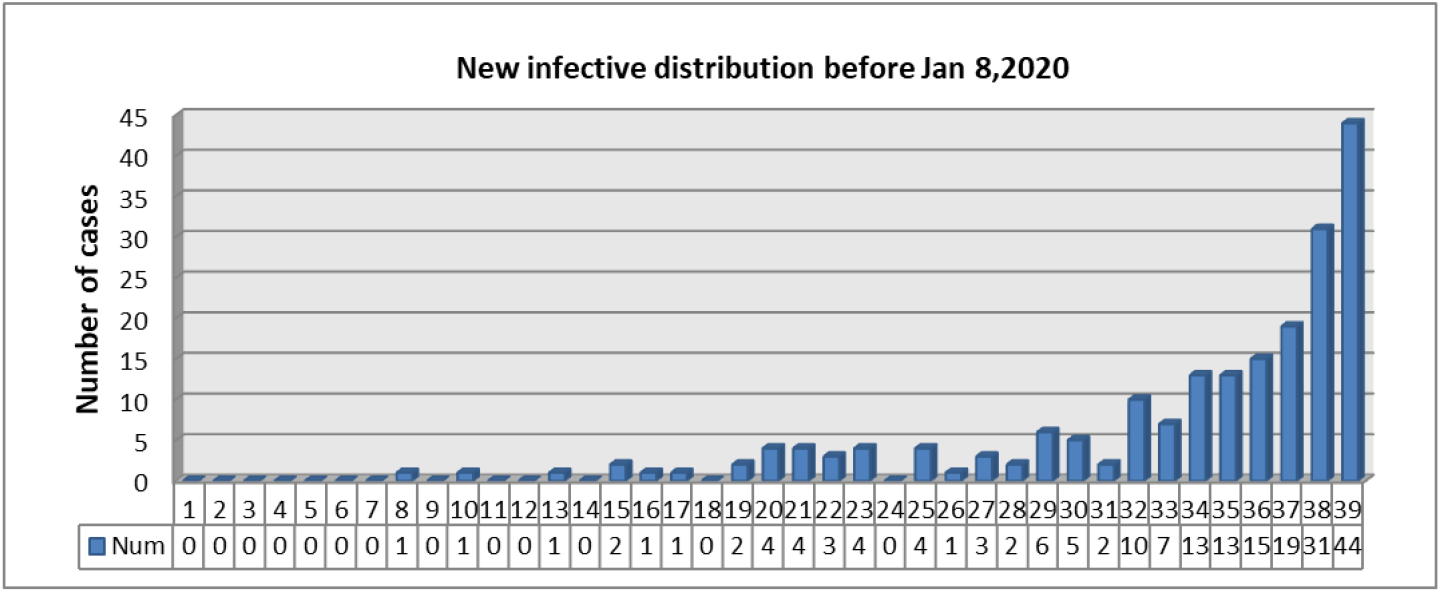
The number of new case every day from Dec 1, 2019, to Jan 8, 2020 [1].

As defined in Equation (6), the number of susceptibles in hospital is equal to the number of infectives stayed in hospital. In fact, *ξI*_*c*(*t*)_ + *I*_*h*(*t*)_ in (6) is substituted by the number of the hospitalized patients in Wuhan city, which is reported by Wuhan Municipal Health Commission (WMHC)[36] and Health Commission of Hubei Province(HCHP)[37]. In addition, the execution of (12) and (13) is a time-consuming process. So, they are reformulated by

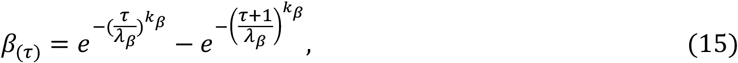

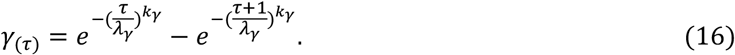

It is readily seen from Equation (10) that the exposed individuals can be classified as belonging to one of *P*_*E*_ + 1 groups. And, the sizes of these groups are changed every day. The update formula is

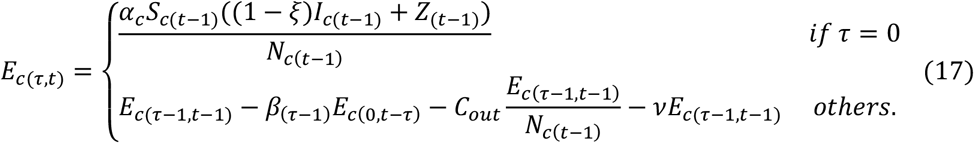

Similarly, there are

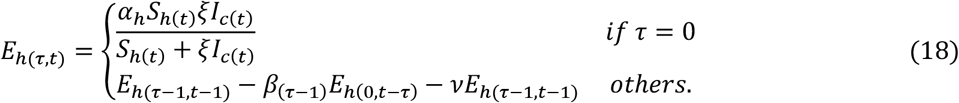

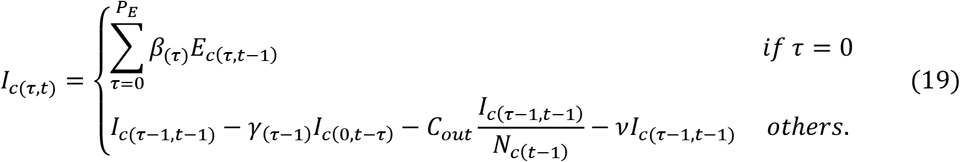

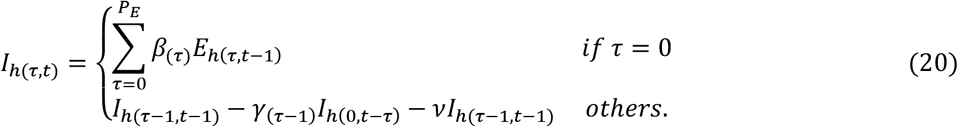

## 5. Solution of parameters

In this section, we propose a solution based on constraint optimization to estimate the parameters of the SEIR-HC model mentioned in Section 4.

### 5.1 Objective function

The epidemic model is devised to estimate the unobserved variables and to predict the transmission process. The output data of the model is desired to be close to the real. Unfortunately, it is difficult for the lack of the credible observations. To alleviate this problem, reasonable data is considered as an alternative. The data collection is completed by integrating multiple data sources:

1. Number of new cases every day from Dec 1, 2019, to Jan 8, 2020, provided by Li et al (in Fig. 3) [1],
2. Number of new cases every day from Feb 20 to Mar 10, 2020, reported by WMHC[36] and HCHP[37],
3. Number of the infectives among the nationals who returned to America[38], Japan[39], South Korea[40] and Singapore[41]from Jan 29 to Jan 31, 2020 (as tabulated in Table I),
4. Number of the infectious hospital staff members is 1102 on Feb 11, 2020, reported by The State Council Information Office of China[42],
5. Numbers of discharged and dead patients reported by WMHC[36] and HCHP[37], sum of which is theoretically equal to the number of the individuals removed by both recovery and death. Note that several numbers were corrected for the contradiction between accumulated cases and new cases.

**Table I.**
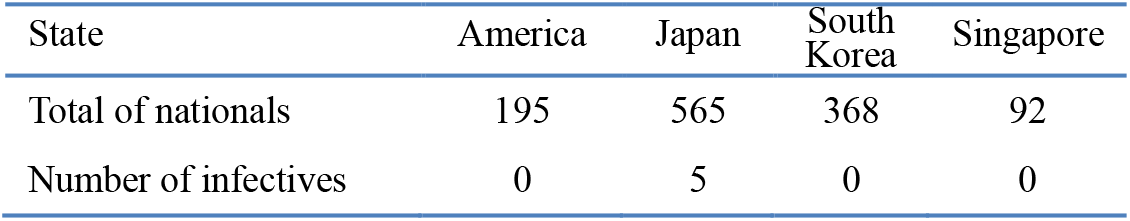
Number of the exposed and infectious individuals among the nationals from Jan 29 to Jan 31.

Based on the above data, the objective function consists of the five error sums of squares as follows:

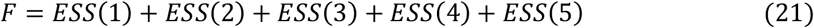

Where

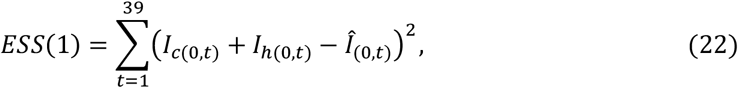

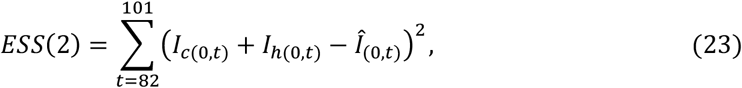

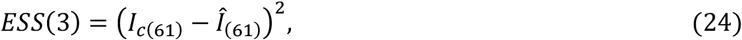

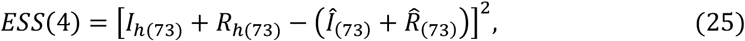

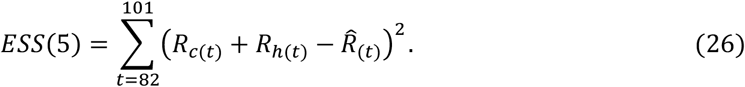

Note that the sign (^) denotes recorded value.

Statistically, the nationals in Wuhan city can be regarded as the samples of population in the community. The ratio of the cases confirmed during the time from Jan 29 to Jan 31 among nationals is approximately equal to that on Jan 30. In this sense, the number of infectives is likely to be about 36880 in Wuhan city on Jan 30.

### 5.2 Initialization of parameters

The parameter estimation is, in general, a complex task partly due to so many unknown parameters: *ξ, ζ, α*_*c*_, *α*_*h*_, *k*_*β*_, *λ*_*β*_, *k*_*γ*_, *λ*_*γ*_. Correct convergence is hard to reach unless a reliable initial guess is provided. Therefore, an optimum initial estimation based on knowledge is crucial in this case. For this reason, all the parameters are dichotomized into two classes. The parameters *ξ, ζ, α*_*c*_, *α*_*h*_ belong to class one and the others class two. Among the first-class parameters, *ξ, ζ* are determined by intervention, *α*_*c*_, *α*_*h*_ reflect the combined effect of virus and control. The second-class parameters are related to the characteristic of the virus.

To estimate the parameters of the first class, we first investigated the implementation of control measures in Wuhan city. From the report of HCHP[37], we found that an absolute increment of the number of confirmed infectives was more than 10 thousand on Feb 12, 2020 compared to about one thousand the day before that day. This implies that many patients are likely to fail to be hospitalized before Feb 12. Taking into account the strict restriction on outdoor activities imposed before, *ξ* and *ζ* are set at 0.8 and 0.2.

The permanent resident population of Wuhan city is about 14 million[34]. Moreover,Wuhan is well known for being the transport hub of China. The contact rate *α*_*c*_ was set 3 for the infectiousness in the community. On the other side, ideally, infections hardly spread in hospital. But, this is not the case for the inadequacy of medical supplies. And, according to the report of HCHP, a lot of them had been distributed after Feb 12. From these points under consideration, the initial contact rate *α*_*h*_ is set equal to 0.05. At the same time, the contact rate is assumed to decrease by 99% after Feb 12.

With regard to class two, the parameters are relevant to the Weibull PDF of the latent period and the infectious period. According to the medical records of 138 patients at Zhongnan Hospital of Wuhan University, the median hospital stay is 10 days[43]. The mean incubation period is 5.2 days[1]. Assuming that the latent period and the infectious period is approximately equal to the incubation period and the length of stay, the mean values of the latent period and the infectious period are preliminarily estimated to be 5 and 10 days, respectively. The profiles of the Weibull PDF with various shape parameters are shown in Fig. 4. As a result, the initial guesses of the parameters *k*_*β*_, *λ*_*β*_, *k*_*γ*_, *λ*_*γ*_ are 2.4, 5.64, 2.4 and 11.28, respectively.

**Fig. 4.**
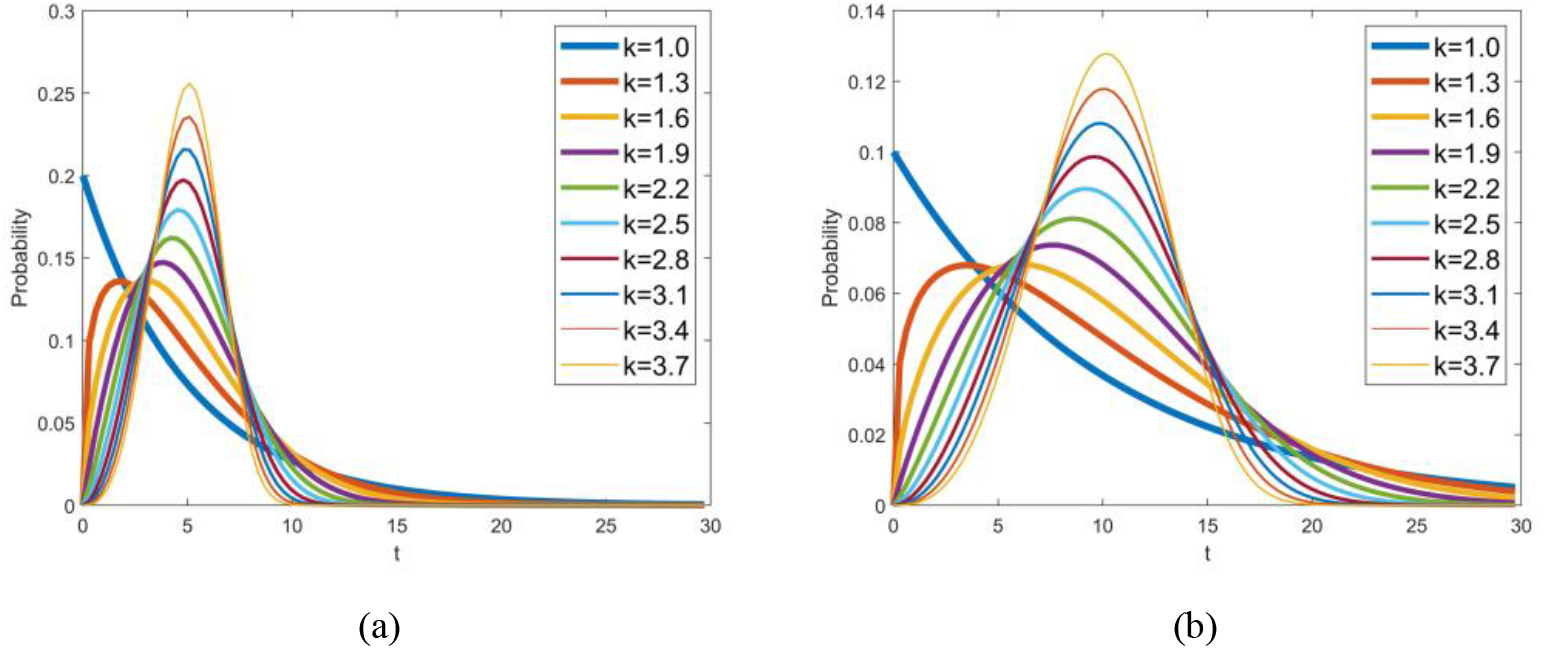
Weibull PDF with various shape parameters while (a) mean value is 5, and (b) mean value is 10.

Similarly, we determined the low bound and upper bound of the parameters. It is worth noting that to cut down the domain may not be a good strategy for global optimization since that may block the way to the global optimal point.

### 5.3 Two-step iterative optimization

Global solutions are usually difficult to locate, whereas the situation may be improved when constraints are add [44]. Consequently, an inequality constraint is defined to allow algorithms to make good choices for search directions. Based on the knowledge that solation and quarantine are a useful control measures[35], the numbers of exposed and infectious individuals in the presence of control efforts are consequentially no more than those in the absence of interventions.

Furthermore, we explored a two-step optimization by adopting the sequential quadratic programming (SQP) method. In theory,if only the mean values of the latent period and the infectious period are unchanged, the result is almost invariable in number. In this sense, for a given mean value, the second-class parameters only affect the shape of the SEIR-HC model. As a result, we searched the optimal solution for the first class by the SQP method at first and take that as the initial guesses of the second class. Then, the second-class parameters are determined by the same way. The above process is iterated many times. The flow chart is presented in Fig. 5.

**Fig. 5.**
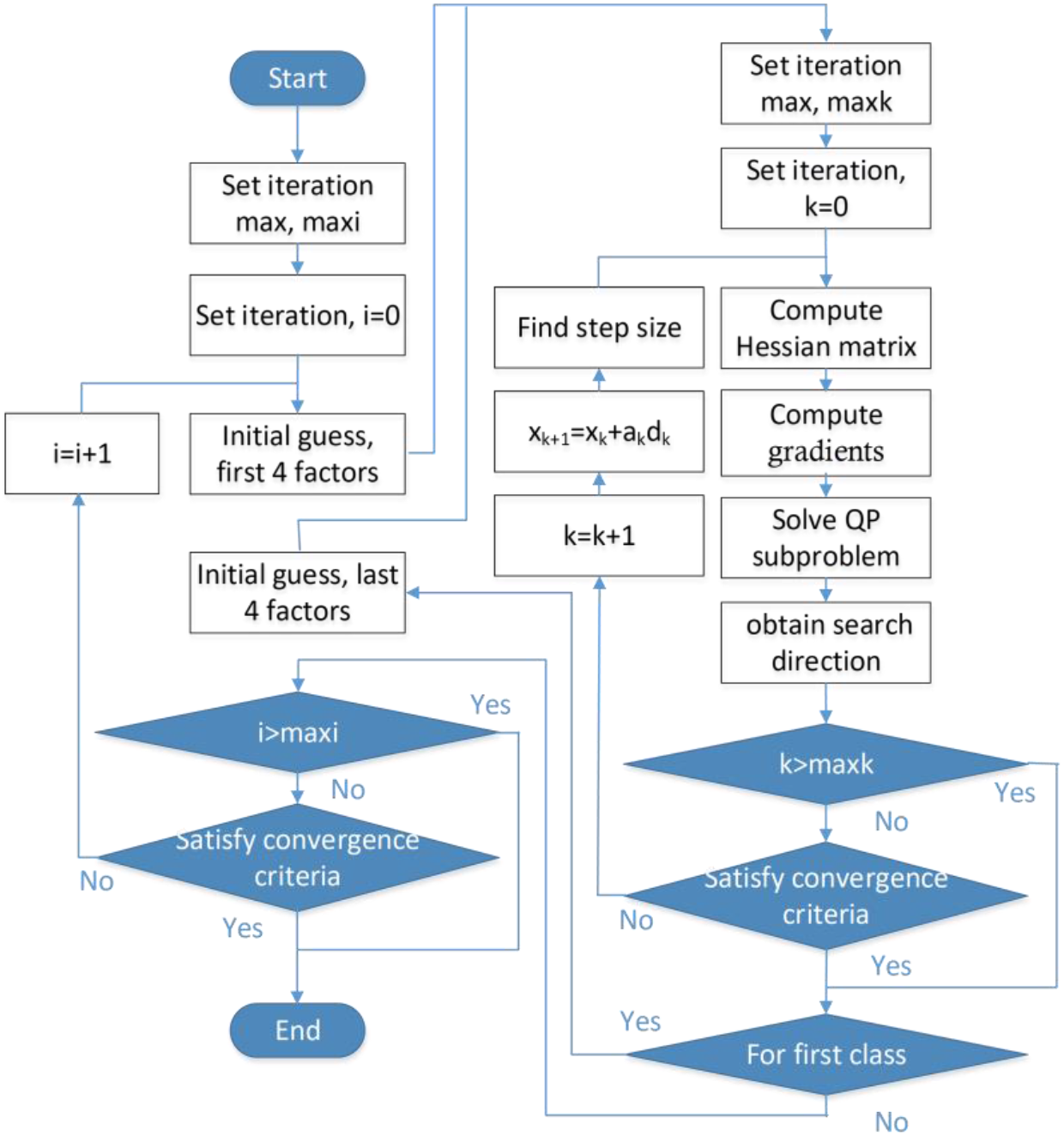
Flow chart of the complete two-step iterative optimization.

In brief, the complete process of the two-step parameter optimization can be divided into four steps:

1. Guess all the parameters and their low bound and upper bound as described in Section 5.2.
2. Estimate the parameters of the first class using the SQP method with the objective function presented in Section 5.1 and inequality constraints stated in this section.
3. Estimate the parameters of the second class in the same manner after the estimated values of the first-class parameters is updated with the result of the second step.
4. Repeat the computing process from (2) to (4) until the bias is small than a given threshold value or cycle index reaches 5.

## 6. Result and discussion

### 6.1 Estimated value of SEIR-HC parameters

After the two-step iterative optimization, all the parameters of SEIR-HC model are determined for COVID-19 in Wuhan. The low bound, the upper bound, the initial value and the estimated results are summarized in Table II. All results fall within the range between the upper and lower limits. Some are close to the initial value, but some far from the guess.

**Table II.** Initial value and bounds of parameters of the SEIR-HC model.

| Parameters | $\xi$ | $\zeta$ | $\alpha_c$ | $\alpha_h$ | $k_\beta$ | $\lambda_\beta$ | $k_\gamma$ | $\lambda_\gamma$ |
| --- | --- | --- | --- | --- | --- | --- | --- | --- |
| Low bound | 0.4 | $10^{-5}$ | $10^{-5}$ | $10^{-5}$ | $10^{-5}$ | $10^{-5}$ | $10^{-5}$ | $10^{-5}$ |
| Upper bound | 0.999 | 0.5 | 16 | 16 | 4 | 30 | 4 | 30 |
| Initial value | 0.8 | 0.2 | 3 | 0.05 | 2.4 | 5.64 | 2.4 | 11.28 |
| Result of step 1 | 0.799 | 0.027 | 2.999 | 0.039 | - | - | - | - |
| Result of step 2 | - | - | - | - | 2.983 | 4.693 | 1.101 | 12.987 |

### 6.2 Epidemiology characteristic of COVID-19

With the parameters estimated, it is easy to derive that the mean and variance of the latent period are 4.19 and 1.53, and those of the infectious period are 12.53 and 11.40. The Weibull PDFs of the latent period and the infectious period are shown in Fig.6. It can be observed that a large proportion of exposed individuals become infectious in a short time and most cases are mild. The cumulative probability of the latent period for 10 days is up to 99.99%, by which the 14-day period of active monitoring is well supported. The difference between the latent period and the incubation period is equal to 1.01 days and the difference between the infectious period and the length of stay is 2.53 days.

It is worth noting that 1 − *ξ*(= 20.1%) infectives, which is equivalent to, on the average, 2.52 days an infective, still stayed in the community before Feb 12. Assuming that the infectiousness is constant during the entire infectious period, the basic reproduction number is up to 7.90 where everyone is susceptible. The basic reproduction number estimated here is compared with others in Table III. Furthermore, even though an infective is hospitalized at the beginning of clinical symptoms, he still can make 3.48 individuals infected. As a result, the outbreak is inevitable in the absence of interventions because of difference between the latent period and the incubation period. Of course, this is not necessary true for districts other than Wuhan city, because basic reproduction number varies according to population density and social enthusiasm besides characteristics of pathogenic bacteria [45].

**Table III.**
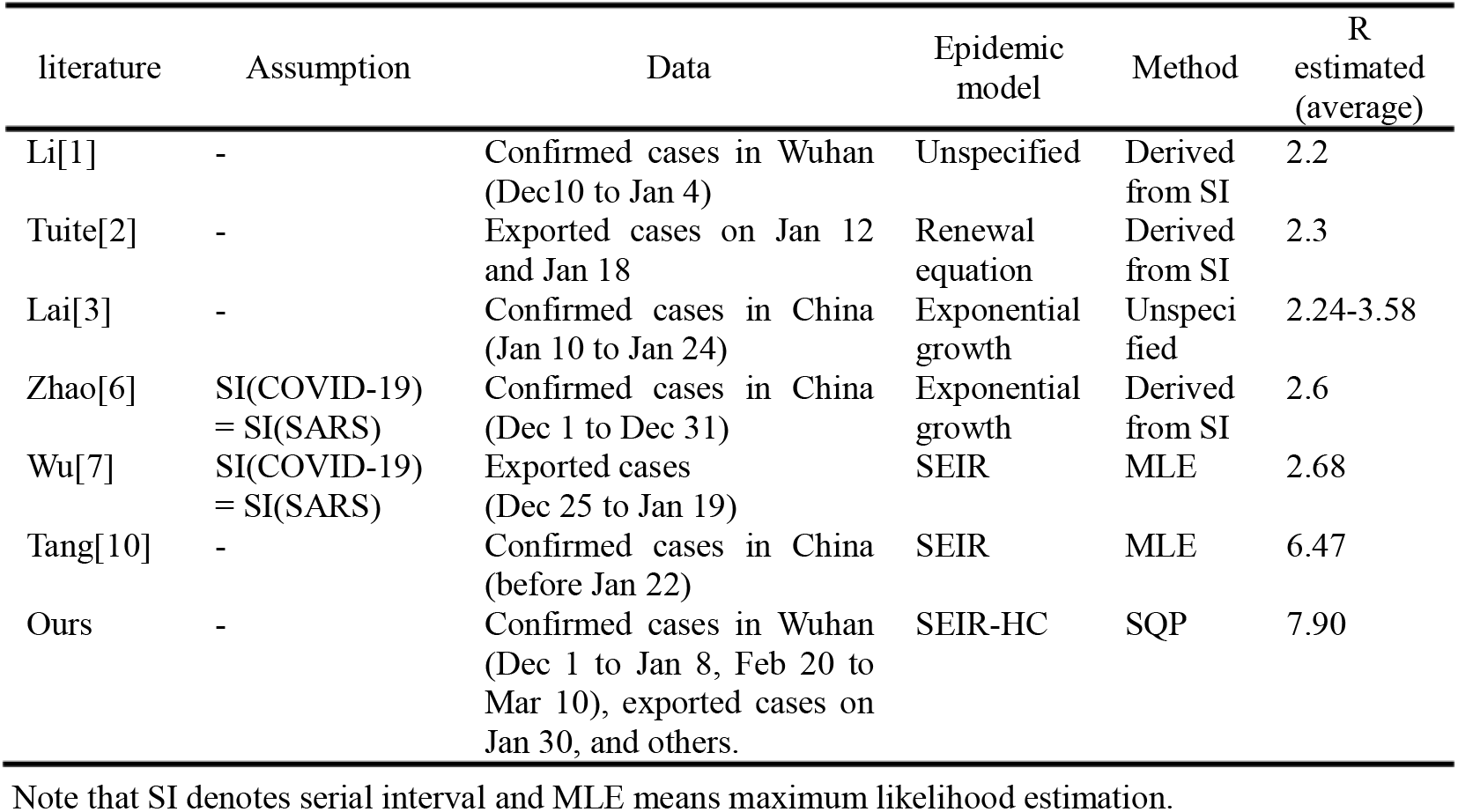
Comparison of basic reproduction numbers.

| literature | Assumption | Data | Epidemic model | Method | R estimated (average) |
| --- | --- | --- | --- | --- | --- |
| Li[1] | - | Confirmed cases in Wuhan (Dec10 to Jan 4) | Unspecified | Derived from SI | 2.2 |
| Tuite[2] | - | Exported cases on Jan 12 and Jan 18 | Renewal equation | Derived from SI | 2.3 |
| Lai[3] | - | Confirmed cases in China (Jan 10 to Jan 24) | Exponential growth | Unspecified | 2.24-3.58 |
| Zhao[6] | SI(COVID-19) = SI(SARS) | Confirmed cases in China (Dec 1 to Dec 31) | Exponential growth | Derived from SI | 2.6 |
| Wu[7] | SI(COVID-19) = SI(SARS) | Exported cases (Dec 25 to Jan 19) | SEIR | MLE | 2.68 |
| Tang[10] | - | Confirmed cases in China (before Jan 22) | SEIR | MLE | 6.47 |
| Ours | - | Confirmed cases in Wuhan (Dec 1 to Jan 8, Feb 20 to Mar 10), exported cases on Jan 30, and others. | SEIR-HC | SQP | 7.90 |

Note that SI denotes serial interval and MLE means maximum likelihood estimation.

### 6.3 Transmission process of COVID-19

With the SEIR-HC model proposed here, the transmission process of COVID-19 epidemic in Wuhan city is reproduced as shown in Fig.7. In our baseline scenario, we estimate that the outbreak would be over before Apr 15, 2020, and the total of infectives no longer increases by Mar 15, 2020. At the same time, the total number of removed individuals would reach 111383 finally (as shown in Fig. 7(a)). Among them, the number of hospital staff members would be up to 2950, which is likely to be slightly more than the reported cases due to asymptomatic infection. The number of the infected individuals in Wuhan on Jan 25, 2020, is estimated to be 46029, which is less than the count 75815 estimated by Wu et al [7]. The number of infectives on Jan 30 is 35962 (2.52% difference to 36880), which is much higher than 20767 (the number of the infected individuals estimated by Nishiura et al [5]). Number of infectious hospital staff members on Feb 11 is equal to 1241 (11.87% difference to 1022).

**Fig. 6.**
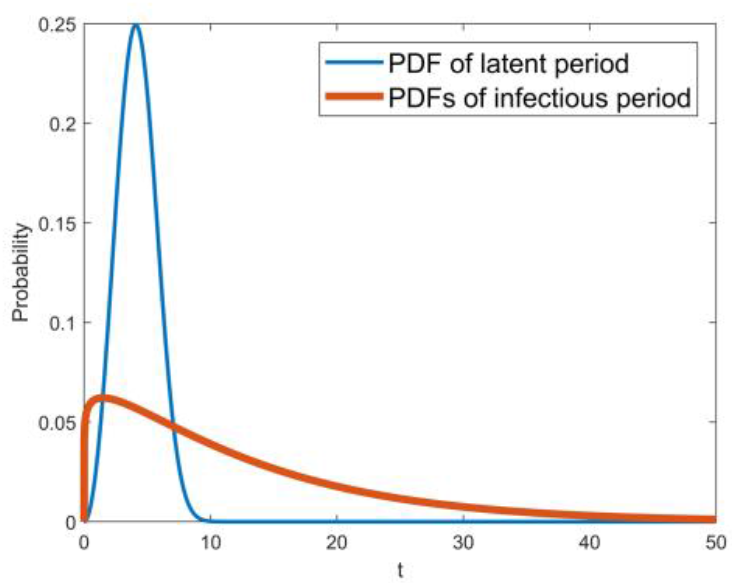
The Weibull PDFs of the latent period and the infectious period.

**Fig. 7.**
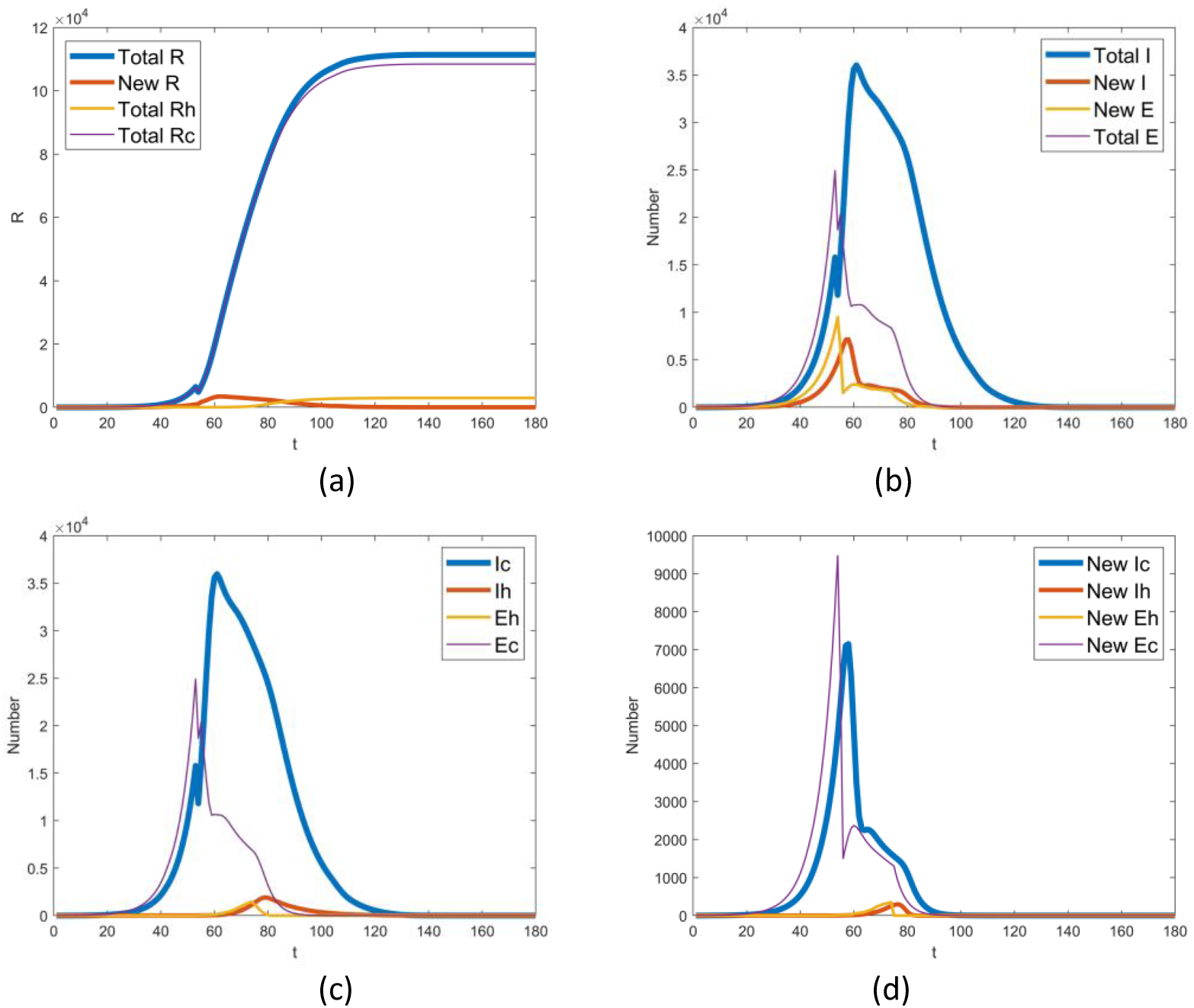
Transmission process of 2019-nCov epidemic in Wuhan city. In the picture, the capital letters E, I, R indicate the exposed, infectious and removed individuals, respectively. And, the lowercase letters c and h mean community and hospital. Note that t indicates the number of days delayed from Dec 1, 2019.

It is readily seen from Fig. 7(b), (c) and (d) that there is a sudden decrease of the number of the exposed individuals on Jan 23 and on Feb 12, which implies control measures launched have a conspicuous effect on the infection rate.

Though mathematical models of epidemic transmission often scarce contrast model output with real-world observations[4], the comparison is necessary to demonstrate the performance of the models and the validity of the results. For this reason, Fig.8 provides a pictorial display of comparison of the estimated data and the reported ones. The two curves show the same trend after Feb 12. But, before that day, the number of infectious individuals estimated here is far more than that reported by WMHC and HCHP. A probable reason for this is the underreporting of incidences before Feb 12[1, 2, 5, 6], which may result in an underestimation of basic reproduction number.

From Fig. 8(a), it seems that there is a delay of the estimated number of the removed individuals relative to the reported. The underlying cause of the delay may be the fact that the discharge time is later than the end time of infectious period for a need for functional recovery. In this sense, it appears to be appropriate that the outbreak in Wuhan terminates later than the expected time. However, the warmer weather is helpful in preventing the virus from reproducing. Given these points, the outbreak is likely to be ended as we expected before if the control measures are kept on as usual. Additionally, it also can be observed from Fig. 8(a) that, in the worst case, up to 33620 infectives failed to be hospitalized on Jan 30.

**Fig. 8.**
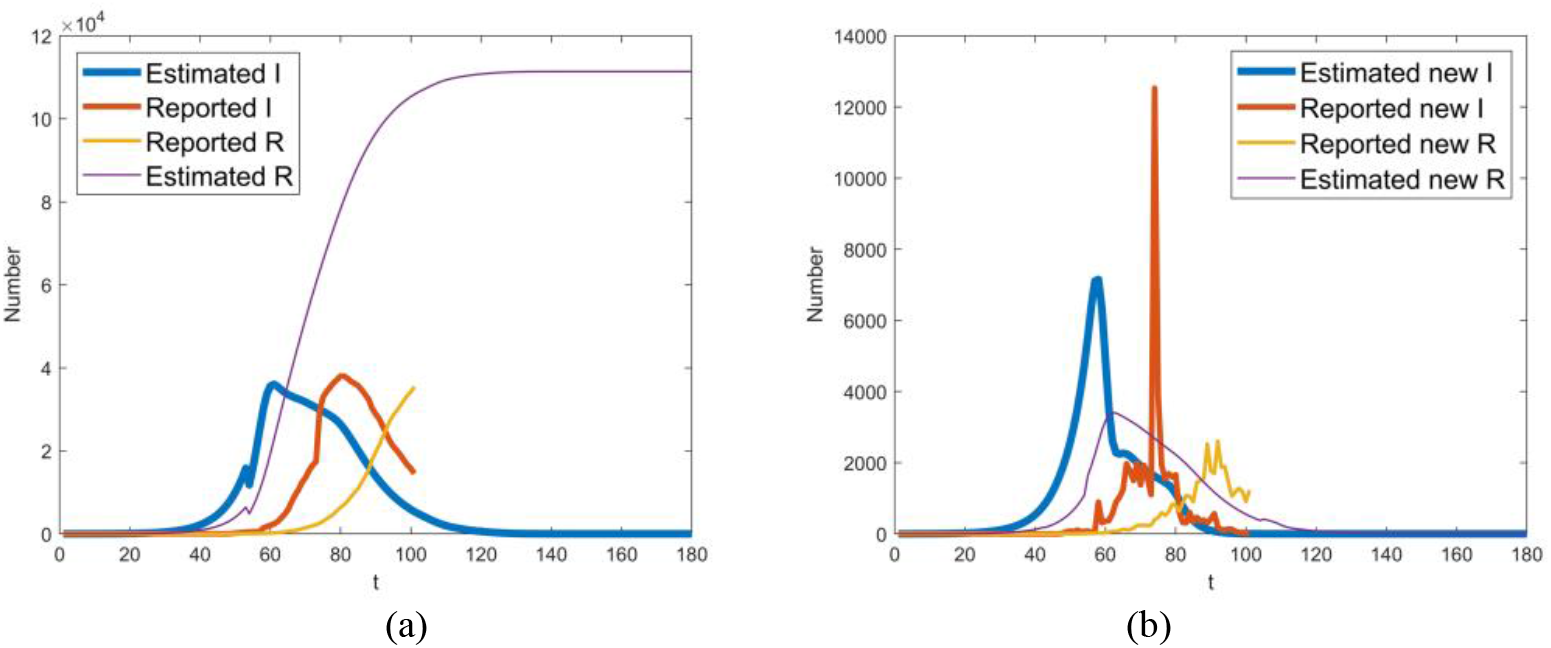
Comparison of the infectious *I* and removed individuals *R* estimated here with the ones reported by WMHC and HCHP. Note that *t* indicates the number of days delayed from Dec 1, 2019.

### 6.4 Control measures

To assess the control measure, a series of experiments are carried out using the SEIR-HC model. The control measures are simulated by the first-class parameters and the effect is captured by the number of infectives.

#### 6.4.1 Effect of control level

Since the proportion of quarantined susceptibles to the total number of susceptibles 1 − *ζ* implies control level, the function of quarantine is tested by adopting *ζ*. From the SEIR-HC model, we can see that *ζ* impacts primarily on the community infection. So, only Ic-t and (Ic+Rc)-t graphs are shown here for space limitations. Fig. 9(a) shows *I*_*c*_varing when *ζ* increases from 0.032 to 0.932 and Fig. 9(b) shows *I*_*c*_ + *R*_*c*_. Fig. 9(c) and (d) display the results with a delay of 0∼9 days. It was clear that the number of infectives dramatically increases with the proportion *ζ* and delay time. As a result, control measures played a key role in prevention of the spread. It also can be found from Fig. 9 that the resulting divergence begins after Jan 28. Therefore, Jan 23, on which the measures imposed, is the right time to stop the outbreak.

**Fig. 9.**
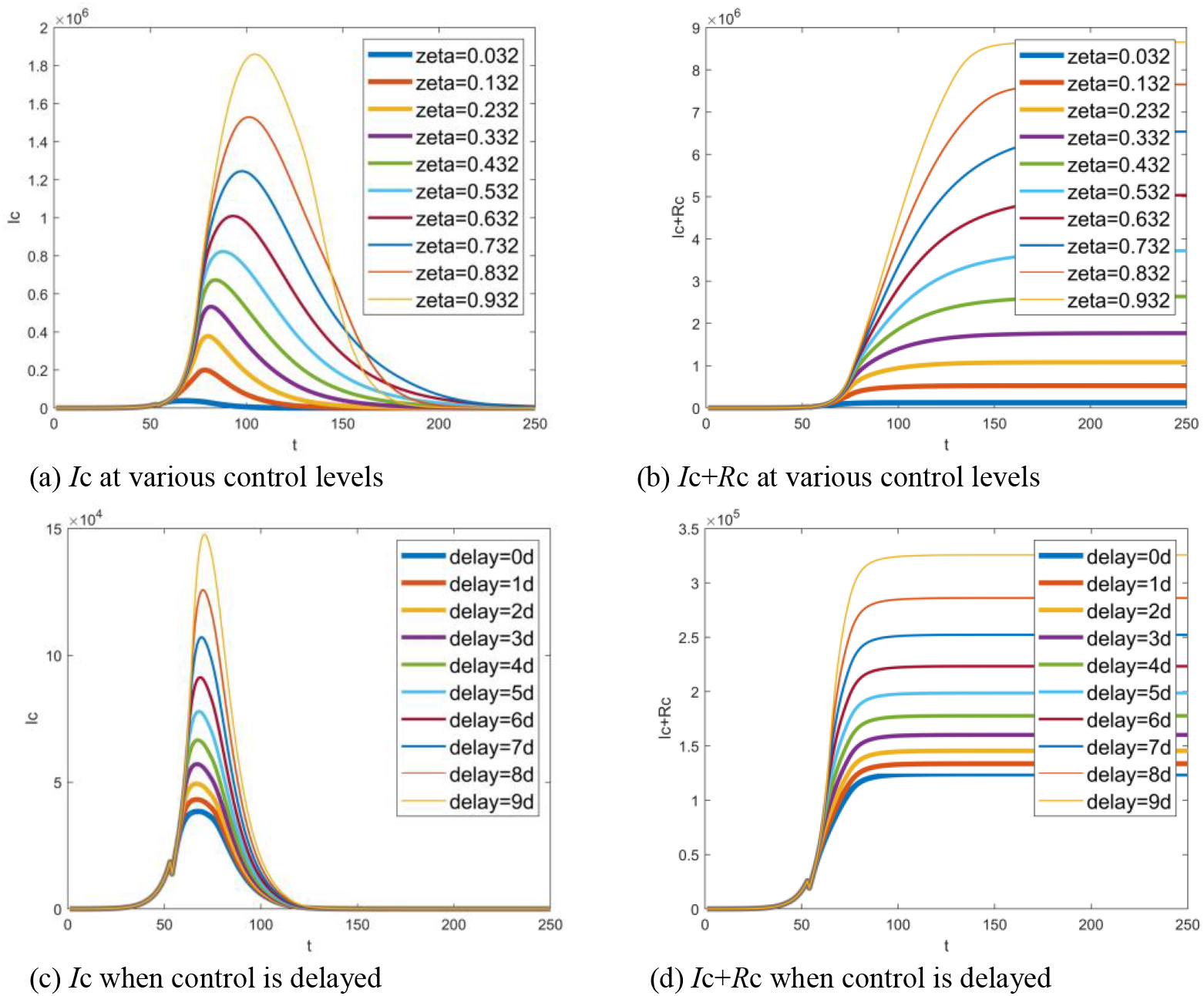
Simulated function of control measures: (a) number of infectives at various control levels, (b) accumulative total of infectives at various control levels, (c) number of infectives when control is delayed, (d) accumulative total of infectives varying with delayed time. Note that *t* indicates the number of days delayed from Dec 1, 2019, while delay=xd means x days delayed from Jan 23, 2020.

#### 6.4.2 Effect of the time of hospital admission

Given the proportion of hospitalized infectives to total number of infectives *ξ*, 1 − *ξ* is equivalent to the proportion of the average time during which infectives stay in the community to the average infectious period, which reflects how quickly the infectives are allowed to hospitalize. In this section, the effect of the time of hospital admission is test by changing parameter *ξ*. Fig. 10(a) shows that the number of infectives in the community varies with *ξ* increase and Fig. 10(b) is the corresponding accumulative value. Fig. 10(c) and (d) depict the case in hospital. The delay of hospital admission makes an increase in the number of infectives both in the community and in hospital. However, the increase of the number of infectives slows down when *ξ* is small enough due to the depletion of susceptibles in the population. It can be observed from Fig. 10(b) that, as a result of a large number of inbound travellers every day, the accumulative total of the infectives can be even more than permanent resident population. In fact, this is almost impossible because the implicit assumption that travel behavior was not affected by epidemic is not valid.

**Fig. 10.**
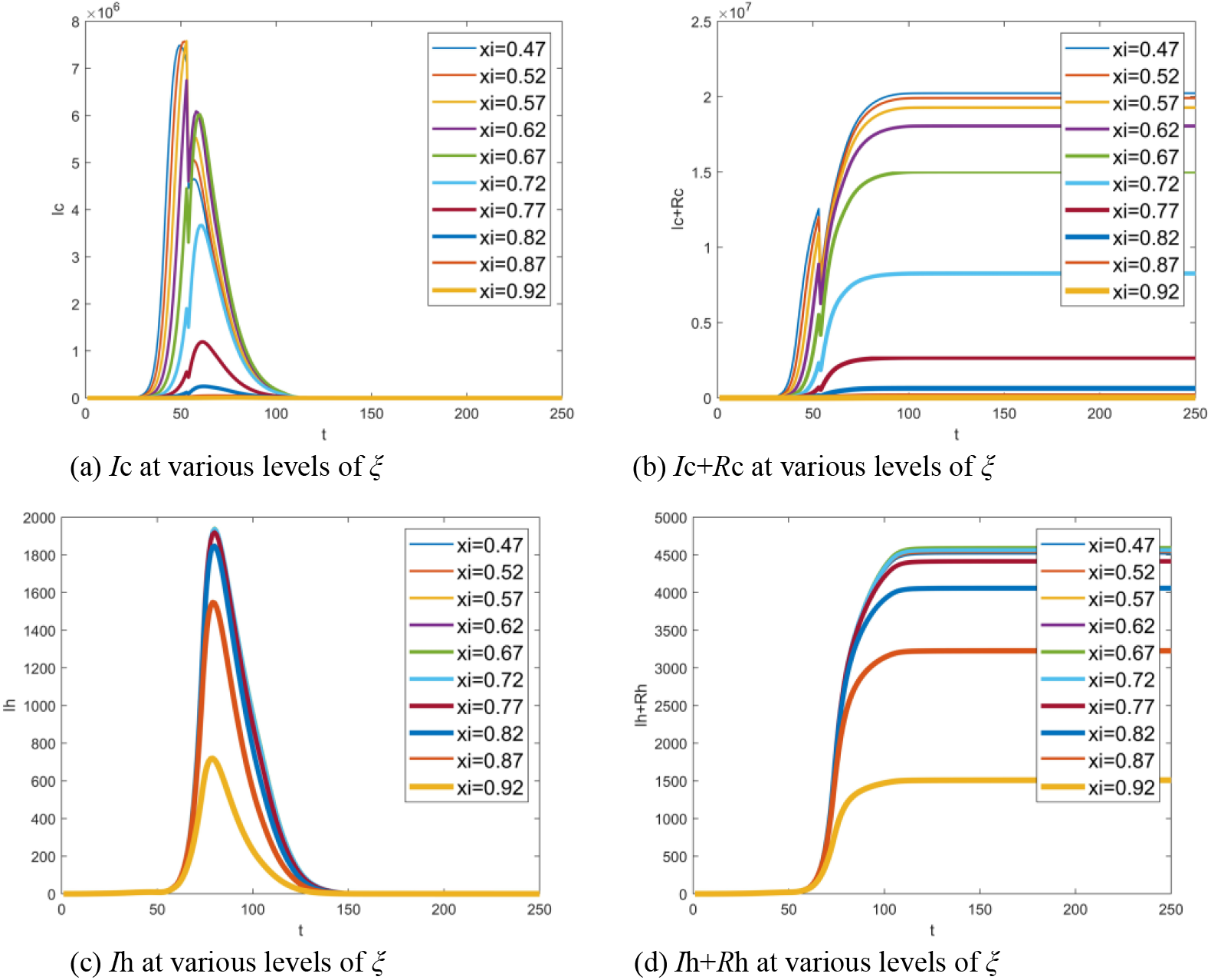
Effect of the time of hospital admission: (a) number of infectives in the community at various proportion *ξ*, (b) accumulative total of infectives in the community at various proportion *ξ*, (c) number of infectives in hospital at various proportion *ξ*, (d) accumulative total of infectives in hospital at various proportion *ξ*. Note that *t* indicates the number of days delayed from Dec 1, 2019.

#### 6.4.3 Effect of contact rate *α*_c_

Contact rate is primarily determined by the nature of pathogen. In fact, it can also be changed in some degree by intervention strategies[23]. Assuming that *α*_c_ is varied from 3.3 to 5.1, the number of infectives in the community varying with *α*_c_ is demonstrated in Fig. 11. A clear result is that the number of infectives in the community increases exponentially with *α*_c_. Note that, following the same idea mentioned in Section 6.4.1, only the number of infectives in the community is shown here.

**Fig. 11.**
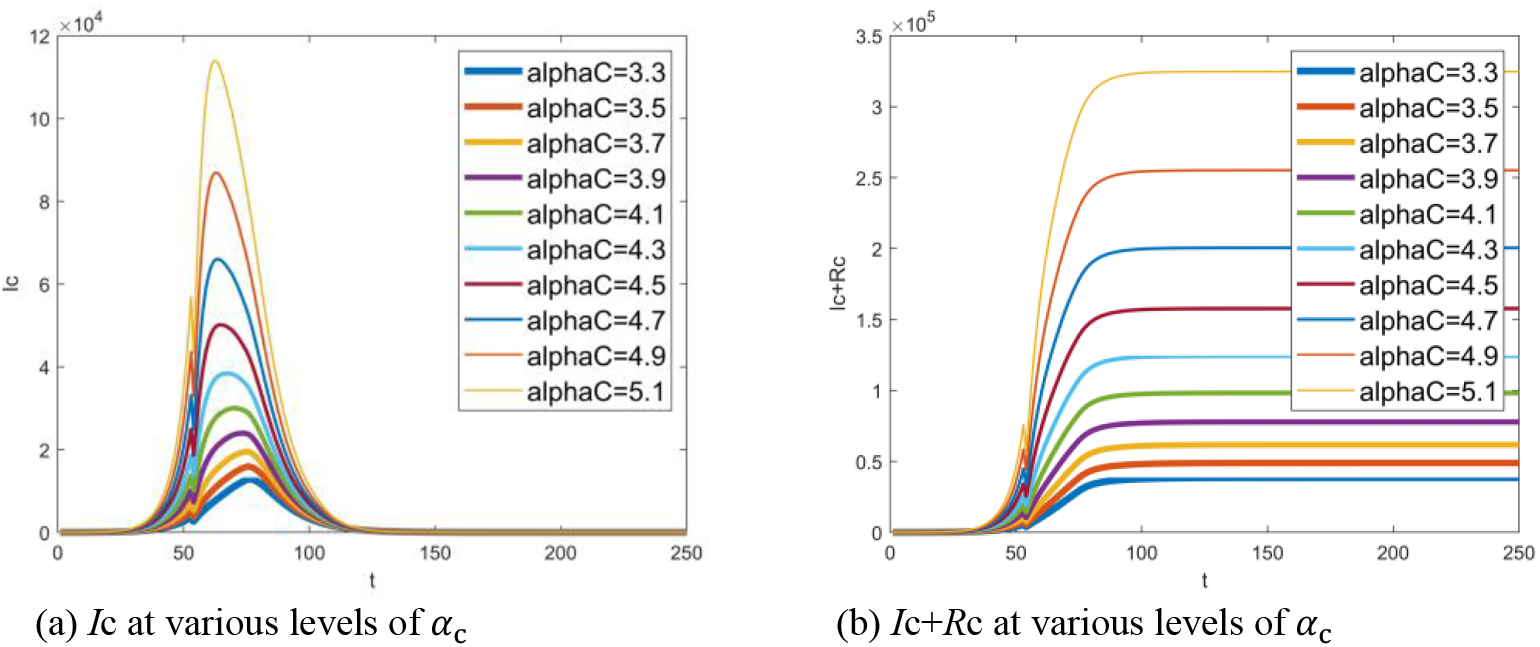
Effect of contact rate *α*_c_: (a) number of infectives in the community at various levels of contact rate *α*_c_, (b) accumulative total of infectives in the community at various levels of contact rate *α*_c_. Note that *t* indicates the number of days delayed from Dec 1, 2019.

**Fig. 12.**
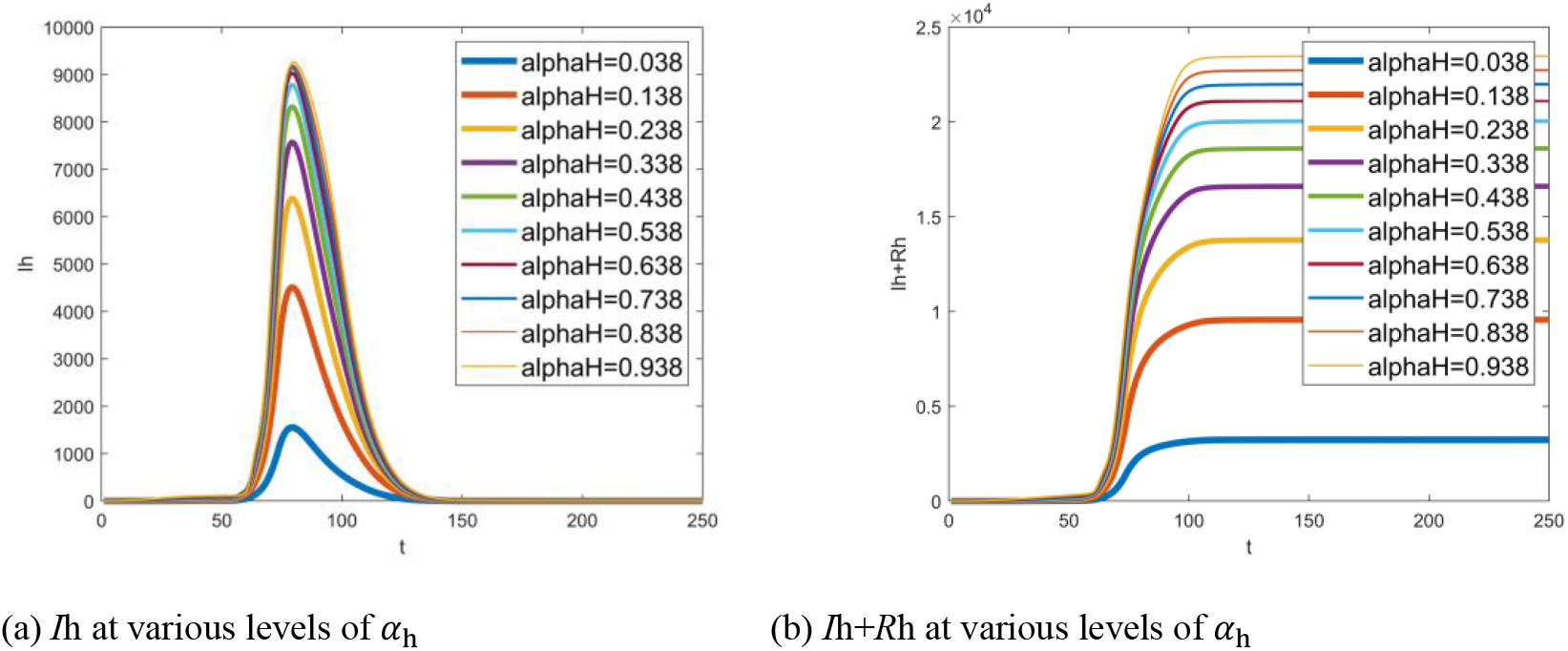
Hospital infections with different contact rate *α*_h_: (a) number of hospital staff members infected by COVID-19 at various levels of contact rate *α*_h_, (b) corresponding cumulated number. Note that *t* indicates the number of days delayed from Dec 1, 2019.

#### 6.4.4 Effect of contact rate *α*_h_

The contact rate *α*_h_ partly reflects the prevention level in hospital. From Fig.12, the number of infectives in hospital increase sharply with *α*_*h*_.

#### 6.4.5 Control methodology of COVID-19

According to the experimental results, it is easy to find out about the control methodology. The basic principle is taking measures as early as possible to lower *ζ, α*_c_, *α*_h_ and to enhance *ξ*. The control measures of COVID-19 list as follows.

1. Keeping in home quarantine and reducing travel to lower *ζ*;
2. Tracing, testing and quarantining the suspected case, immediately isolating symptomatic individuals and speeding hospital admission to enhance *ξ*;
3. Strengthening personal protection to lessen *α*_c_ and *α*_h_.

## 7. Conclusion

This work provides the SEIR-HC model, a novel SEIR model with two different social circles, for formulating the transmission dynamics of COVID-19, and a two-step optimization method exclusively designed for parameter estimation of SEIR-HC model. With the model, the spread process of COVID-19 is reproduced clearly even without enough observation data. The latent period, infectious period and basic reproduction number of COVID-19 are estimated to be 4.19, 12.53 and 7.90, respectively. Obviously, COVID-19 is highly transmissible. The outbreak in Wuhan is anticipated to be over before Apr 15, 2020. The total number of removed individuals would reach 111383 finally. Among them, the number of hospital staff members would be up to 2950.

According to the SEIR-HC model, the principle of prevention and control of COVID-19 is taking measures as early as possible to lower *ζ, α*_c_, *α*_h_ and to enhance *ξ*. A set of measures such as quarantine have significant impacts on lessening the spread. By the way, an international effort is required to prevent virus transmission since COVID-19 has spread all over the world.

As a whole, the conclusions are well interpretable and reasonable. As evidenced by the success in estimation and prediction, the SEIR-HC model is useful. Although the results are based on the data from Wuhan and hence they are not necessary to be reliable for other cities, the SEIR-HC model is valid everywhere, which allows us to capitalize on new data streams and lead to an ever-greater ability to generate robust insight and collectively shape successful local and global public health policy.

## Data Availability

(1) Notification on pneumonia of the new coronavirus infection reported by Wuhan health committee. 2020 [cited 2020 Mar 10]; Available from: http://wjw.wuhan.gov.cn/front/web/showDetail/2020012009077.
(2) Bulletin of hubei provincial health committee on pneumonia caused by novel coronavirus. 2020 Mar 10, 2020 [cited 2020 Mar 10]; Available from: http://wjw.hubei.gov.cn/fbjd/tzgg/202001/t20200121_2013873.shtml.
(3) Quarantine of evacuees at march air reserve base ends. 2020 Feb 11, 2020 [cited 2020 Mar 10]; Available from: https://nbcpalmsprings.com/2020/02/11/quarantine-of-evacuees-at-march-air-reserve-base-ends/.
(4) Two new cases of asymptomatic infection in Japan were the third group of people evacuated from wuhan to Japan. 2020 Feb 3, 2020 [cited 2020 Mar 10]; Available from: https://m.chinanews.com/wap/detail/zw/hm_rbxhqb/2020/02-03/hm56146.shtml.
(5) Details: a seventh case of novel coronavirus infection has been confirmed in the republic of Korea. 2020 Jan 31, 2020 [cited 2020 Mar 10]; Available from: https://cn.yna.co.kr/view/ACK20200131002500881.
(6) Past updates on COVID-19 local situation. 2020 Mar 10, 2020 [cited 2020 Mar 10]; Available from: https://www.moh.gov.sg/covid-19/past-updates.
(7) Three departments: a number of measures to care for anti-epidemic frontline medical staff. 2020 Feb 15, 2020 [cited 2020 Mar 10]; Available from: http://www.gov.cn/xinwen/2020-02/15/content_5479035.htm.

## Acknowledgment

This work was supported by National social science foundation of China (16BXW005), Chongqing Basic Science and Frontier Research Project, China (No. cstc2017jcyjAX0007, cstc2017jcyjAX0386), Project Foundation of Chongqing Municipal Education Committee, China (Grant No. 17SKG050), and Sichuan Provincial Key laboratory of Robot Technology Used for Special Environment (Southwest University of Science and Technology), China (Grant No. 17kftk02).

## Notes

### Competing Interest Statement

The authors have declared no competing interest.

